# Seroprevalence of viral hemorrhagic fevers and arboviral infections among febrile patients in Forest Guinea

**DOI:** 10.64898/2026.09.02.26362016

**Authors:** Fara Raymond Koundouno, Youssouf Sidibe, Hugo Soubrier, Karifa Kourouma, Soua Koulemou, Tamba Elie Millimouno, Fernand M’Bemba Tolno, Saa Lucien Millimono, Kékoura Ifono, Faya Moriba Kamano, Mamadou Dioulde Barry, Bely Sonomy, Mariame Traore, Kaba Keïta, Mette Hinrichs, Beate Becker-Ziaja, Emily Victoria Nelson, Christine Jacobsen, Anke Thielebein, Lisa Oestereich, Meike Pahlmann, Beatriz Escudero-Pérez, Ralf Krumkamp, Stephan Günther, Moussa Kolié, Kaba Keïta, Sira Hélène Guilavogui, N’Faly Magassouba, Sanaba Boumbaly, Giuditta Annibaldis, Sophie Duraffour

## Abstract

Between 2017 and 2024, routine diagnostics surveillance in Guinea detected several confirmed viral hemorrhagic fever (VHF) cases among febrile patients. To assess exposure to Lassa virus and other viral pathogens, we conducted a retrospective cross-sectional seroprevalence study in Forest Guinea (Guéckédou and N’Zérékoré). Lassa virus seroprevalence was 56.0% in the Guéckédou study group and 29.8% in the N’Zérékoré study group. Seropositivity increased with age in both study groups, suggesting cumulative exposure. Samples from Guéckédou laboratory were also tested for other pathogens, and antibodies against Marburg virus (5.8%), Zika virus (5.4%) and Crimean-Congo haemorrhagic fever virus (1.2%) were detected.

## Background

Viral hemorrhagic fevers (VHFs) and arboviral diseases continue to pose a major public health challenge in low- and middle-income countries (LMICs) with limited diagnostic capacity. In Guinea, substantial efforts supported the establishment of specialized decentralized laboratory capacities in Forest Guinea after the 2014–2016 Ebola virus disease (EVD) epidemic and the 2021 resurgence. Notably, two laboratories in Guéckédou and N’Zérékoré have been key in strengthening regional surveillance for filoviruses and Lassa virus (LASV), enabling timely detection of these life-threatening pathogens (1–3). While routine diagnostics are limited to a small set of relevant high-consequence pathogens, gaps in the local surveillance systems still remain, with a large number of undiagnosed febrile illnesses (3). In this context, retrospective serological studies can help uncovering pathogen circulation in the region, thereby informing national health policy and surveillance strategies. As such, retrospective studies in Guinea have already identified a broad range of viral and bacterial pathogens among cases of acute febrile illness, highlighting the complexity of pathogen circulation and the need for improved laboratory-based differential diagnosis (4, 5). In Sierra Leone, a laboratory-based retrospective sero-epidemiological approach revealed the co-circulation of diverse pathogens, including LASV and Marburg virus (MARV), in undiagnosed or subclinical infections (6).

Guinea is considered endemic for Lassa fever (LF). However, the limited number of reported acute LF cases (approximately 6 per year) raises questions about the extent to which LF is recognised and diagnosed (3). LASV circulation in Guinea has mainly been inferred from cross-sectional serological studies in communities in the upper and forested regions, which consistently showed evidence of LASV exposure as indicated by anti-LASV immunoglobulin G (IgG) seroprevalence rates ranging from 59.6% to 82.3% (7, 8). Similarly, data from neighboring communities in Sierra Leone (9) and Liberia (10) have further highlighted substantial LASV circulation across the Mano River Union region bordering Forest Guinea, with IgG seroprevalence rates of 16.0% and 45.0%, respectively. However, data on LF among febrile patients in Forest Guinea remain limited, with only one study conducted in the 1990s involving five government hospitals as sentinel surveillance sites (11). While EVD serosurvey studies have been conducted (12), data on potential MARV exposure remain lacking despite its emergence in 2021 (2) and ecological evidence of MARV-positive bats in Forest Guinea (13). In parallel, arboviruses have emerged as an increasingly important group of pathogens in the region, with growing evidence of their active circulation, including one documented Zika virus (ZIKV) case in upper Guinea, and the detection of Crimean-Congo hemorrhagic fever virus (CCHFV) in ticks in middle Guinea (14), yet no documented serosurvey data exist among febrile individuals.

To address these gaps, serological laboratory capacity was established in Forest Guinea at two laboratories in Guéckédou and N’Zérékoré. The aim of our study was to better characterize the potential hidden burden of viral infections in the area.

## Methods

### Ethics Statement

This research has been approved by the National Ethics Committee of Guinea (CNERS), numbers 197/CNERS/25 and 070/LRE/CNERS/26.

### Study design and study group

This study employed a laboratory-based retrospective cross-sectional design. Leftover diagnostic samples collected, tested, and stored at the *Laboratoire des Fièvres Hémorragiques Virales de Guéckédou* (LFHV-GKD), Guéckédou, and at the *Laboratoire des Fièvres Hémorragiques Virales de l’Hôpital Régional de N’Zérékoré* (LFHV-HRNZE), N’Zérékoré, as part of routine laboratory diagnostic investigations of suspected VHF cases (3), were randomly selected for serology testing. The diagnostic database was filtered to identify approximately 500 eligible samples per study group. Samples were included if they were: (i) collected between the 1 January 2017 and 31 December 2021, for LFHV-GKD, or from 1 February 2021 until the 31 December 2024 for LFHV-HRNZE; (ii) identified as plasma derived from whole-blood; (iii) negative by real-time reverse transcription polymerase chain reaction (RT-PCR) testing for *Orthoebolavirus* and or *Orthomarburgvirus*, and or LASV, and or yellow fever virus and or dengue virus; (iv) having availability of demographic information, with no more than one missing variable in sex, age, profession, or residence. Samples were excluded if (i) they were positive for at least one VHF pathogen as tested by RT-PCR, (ii) had insufficient volume (less than 0.5 mL), or (iii) had inadequate quality (e.g., hemolyzed specimen). Basic demographic data were included in the laboratory database, which was curated prior to statistical analysis of the final cleaned dataset using Stata (v.18; StataCorp, USA).

### Serology testing

Prior to testing, all plasma samples were inactivated in a high-biosafety containment cabinet with Triton-X 100 at a final concentration of 1% in phosphate-buffered saline (PBS). Commercially available Enzyme-Linked Immunosorbent Assay (ELISA) kits were used according to the manufacturer’s instructions. For LASV, CCHFV, and ZIKV, assays used included the BLACKBOX^®^ LASV (nucleoprotein or NP) IgG ELISA, the BLACKBOX^®^ CCHFV (NP) IgG ELISA, and the BLACKBOX^®^ ZIKV (ED3) IgG ELISA kits (manufactured by Diagnostics Development Laboratory (DDL) before 2023, and by Panadea Diagnostics thereafter; Hamburg, Germany) (Supplementary file). The ZIKV (ED3) IgG ELISA assay included flaviviruses competitor molecules to improve specificity by reducing cross-reactive antibody binding. For MARV, the Qualitative Human Marburg Virus Antibody (GP) IgG ELISA kit (MBS109415) from MyBioSource (San Diego, USA) was used (Supplementary file).

## Results

### Study population

A total of 989 samples were selected across two laboratory-based study groups at LFHV-GKD (n=512) and LFHV-HRNZE (n=477). The median age was 35 years (IQR: 25–50; range: 2– 85) in LFHV-GKD and 36 years (IQR: 19–53; range: 1–95) in LFHV-HRNZE (Table S1). Younger individuals (<20 years) were slightly less represented in the LFHV-GKD group than in LFHV-HRNZE (12.5% vs 25.6%, respectively). In line with this, occupational distribution differed between the two sites, with a higher proportion of students (including pre-school-aged children) in LFHV-HRNZE (Table S1). Sex distribution was balanced in both study groups. Participants predominantly originated from Guinea (99.3%), specifically from N’Zérékoré administrative region (Table S1). At the prefecture level, differences in geographic distribution mirrored the laboratory locations, with the LFHV-GKD group originating mainly from Guéckédou prefecture (95.3%), and the LFHV-HRNZE group from N’Zérékoré prefecture (81.1%), as well as from the neighboring prefectures of Macenta, Beyla, Lola and Yomou.

### LASV anti-NP IgG seroprevalence in febrile cases in both study groups

Prevalence differed between the two study groups, with 56.0% of the samples tested at LFHV-GKD positive for IgG LASV anti-NP, compared with 29.8% at LFHV-HRNZE (Figure 1A and Table S2). While the analysis was stratified by the available demographic characteristics, both sexes showed similar seroprevalence within each group (Table S2). Domestic and farming workers had the highest positivity rates in the LFHV-GKD group (61.6% and 73.4%, respectively), while all occupations showed similar seroprevalence in the LFHV-HRNZE group (30-38%), except for students (18.7%) (Table S2). Generally, younger individuals (<20 years) showed a lower positivity rate than older age groups at both sites, with the LFHV-HRNZE group showing a considerably lower prevalence (16.4% vs 32.8%) in the LFHV-GKD group) (Table S2). Interestingly, while the percentage of anti-LASV IgG positivity in the LFHV-GKD increased steadily with age, in the LFHV-HRNZE it increased up to age 20 and plateaued thereafter (Figure 1B and Table S2). Analysis by patient’s provenance did not provide substantial insight, as the study population appeared to reflect the geographical distribution across prefectures (i.e., Guéckédou and N’Zérékoré) reasonably well; results for the other prefectures must be interpreted with caution because of limited sample sizes (Table S2).

**Figure 1.**
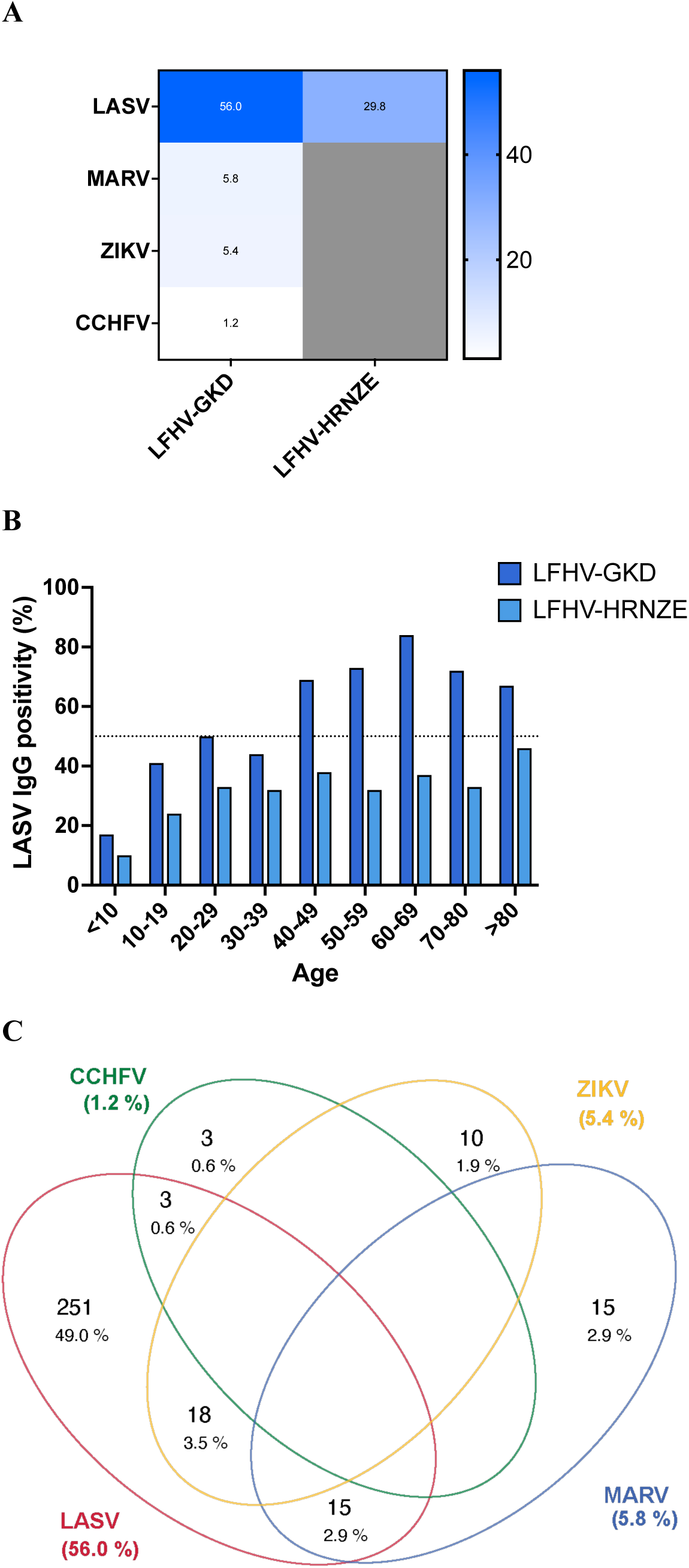
Seroprevalence of IgG antibodies against viral pathogens in febrile cohorts from each laboratory. (A) Heat map showing IgG seroprevalence for each tested pathogen by cohort (LFHV-GKD, Guéckédou laboratory; LFHV-HRNZE, N’Zérékoré laboratory). Darker blue indicates higher seroprevalence; grey indicates pathogens not tested in a given cohort. LASV, Lassa virus; MARV, Marburg virus; ZIKV, Zika virus; CCHFV, Crimean-Congo haemorrhagic fever virus. (B) Age-stratified IgG seroprevalence against LASV by 10-year age groups in each cohort. Colours correspond to cohorts as indicated in the legend. The dotted horizontal line indicates 50% seropositivity. (C) Venn diagram illustrating dual seroreactivity to LASV (red), ZIKV (yellow), MARV (blue), and CCHFV (green) in cohort LFHV-GKD. If no numbers are presented, no cases are observed in the respective groups. Percentages are calculated among all tested individuals (n=512).

### Seroprevalence of other pathogens in febrile cases in the Guéckédou study group

Additionally, the LFHV-GKD study group was tested for antibodies (IgG) against MARV, ZIKV, and CCHFV. Overall seroprevalence was low for all three pathogens: 5.8% for MARV, 5.4% for ZIKV, and 1.2% for CCHFV (Figure 1A and Table S3). Stratified descriptive analyses by age group, sex, geographic provenance, and occupation showed no striking differences across demographic characteristics (Table S3). Specifically, no evidence of increasing seropositivity with age was observed for these pathogens, unlike with LASV (Table S3).

### Co-seropositivity

Co-seropositivity analysis indicated minimal overlap among pathogens, with 54.5% of the samples tested positive for only one pathogen and 7.0% for two, while no samples showed triple or greater reactivity (Table S4). Dual sero-reactivity always involved LASV in combination with another pathogen, with a prevalence of (i) 2.9% for the LASV-MARV combination, (ii) 3.5% for the LASV-ZIKV combination, and (iii) 0.6% for the LASV-CCHFV combination (Figure 1C).

## Discussion

Our study provides evidence of different range of exposure to LASV, MARV, ZIKV, and CCHFV among febrile patients in the study area, using residual diagnostic samples. Routine lab testing has reported a VHFs real-time RT-PCR positivity rate of about 1% among febrile cases (3), but our findings suggest that this figure may not accurately reflect the extent of pathogens’ circulation in the region. LASV anti-NP IgG was of 56.0% in the LFHV-GKD study group and 29.8% in the LFHV-HRNZE study group, indicating that exposure to the virus has occurred far more frequently than would be inferred from reported clinical cases (3), suggesting substantial under-detection of infections, including mild or asymptomatic cases. Although differences in prevalence between the two groups should be interpreted with caution given the cross-sectional, laboratory-based design and differing study periods, the findings provide clues about local transmission dynamics. The association between increasing age and LASV seropositivity, coupled with lower seroprevalence among younger individuals, may be consistent with cumulative exposure over time. However, the age-specific trends differed between the two study groups, suggesting that transmission may not occur uniformly across the two areas. Nevertheless, differences in age distribution between the laboratory-based groups may have influenced the observed patterns.

These observations are consistent with evidence of sustained LASV circulation in both areas. Previous investigations have documented LASV-infected rodent reservoirs in N’Zérékoré city (15), while LF-confirmed human cases have been reported from the study areas in recent years (3). In addition, genomic surveillance data from N’Zérékoré prefecture suggest the circulation of at least two distinct LASV subgroups, one related to strains circulating in Guinea and another linked to strains from neighboring Liberia (16). Together, these findings emphasize the need for future ecological and longitudinal studies to better understand how environmental, demographic, and behavioural factors shape LASV transmission dynamics in both areas. Beyond LASV, the detection of IgG antibodies against MARV, ZIKV, and CCHFV in the LFHV-GKD group highlights the diversity of viral pathogens to which populations in Forest Guinea are exposed. Although these pathogens were less considered in the routine evaluation of febrile illness in recent years, our results suggest that they may contribute to a proportion of undifferentiated fever cases. Interestingly, age-stratified seroprevalence patterns for these viruses did not show the same progressive increase observed for LASV, potentially reflecting more episodic spillover events rather than continuous exposure. Although MARV seroprevalence was low (5.8%), this study provides, to our knowledge, the first serological evidence of MARV exposure in Guinea and establishes a baseline for future assessments, with estimates comparable to those reported previously in Sierra Leone (10.7%) (6). While data on the distribution of arboviral diseases remain limited in Guinea, evidence of ZIKV exposure (5.4%) is particularly notable given its potential consequences for maternal and neonatal health. The low CCHFV seroprevalence observed (1.2%) is consistent with previous reports from 2016–2019 (14), suggesting a similar pattern of limited exposure. These serological findings still highlight the need to strengthen awareness of arboviral circulation in the region, particularly among clinicians, to improve recognition of suspected cases and promote diagnostic testing.

Several limitations should be considered when interpreting these findings. Samples were not exclusively derived from a single geographical area or time period, and financial constraints prevented testing of the full pathogen panel across both study groups. However, this study demonstrates the feasibility and value of leveraging existing decentralized laboratory capacity for serological investigations in Guinea. Serological studies can provide additional information on pathogen exposure and circulation, helping identify underdiagnosed infections, reveal gaps between community exposure and clinically detected disease, and inform future research, monitoring, preparedness, and public health responses.

## Data Availability

All data produced in the present study are available upon reasonable request to the authors.

## Acknowledgments

We thank the Ministry of Health of Guinea for their support and facilitation of this study. We also thank the health workers involved in patient care and sample referral, which made this work possible. We gratefully acknowledge Petra Emmerich and Ronald von Possel for their valuable technical contributions and support throughout the project.

## Author contributions

Conceived and designed the study: F.R.K., Y.S., H.S., N.M., S.B., G.A., S.D.

Collected data and/or performed laboratory testing: F.R.K., Y.S., H.S., Kar.K., S.K., T.E.M., F.M.T., S.L.M., K.I., F.M.K., M.D.B., B.S., M.T., Kab.K. (Mr.), M.H., B.B.Z., G.A.

Data analysis: H.S., R.K., G.A.

Project implementation: F.R.K., Y.S., H.S., M.H., B.B.Z., E.V.N., C.J., A.T., L.O., M.P., B.E.P., S.G., M. K., Kab.K. (Dr), S.H.G., N.F.M., S.B., G.A., S.D.

Funding acquisition: M.P., S.G., N.M., S.B., S.D.

Wrote the manuscript: F.R.K., Y.S., H.S., G.A., S.D. Edited the manuscript: all authors.

All authors read and approved the contents of the manuscript.

## Declaration of generative AI and AI-assisted technologies in the writing process

During the preparation of this work, the author(s) used ChatGPT / free version in order to edit some sentences. After using this tool/service, the author(s) reviewed and edited the content as needed and take(s) full responsibility for the content of the publication.

## Supplementary file

### Methods

#### Serological assays and assay validation

The serological assays used in this study have been previously developed and validated or independently evaluated for the detection of virus-specific IgG antibodies. The BLACKBOX® LASV and CCHFV NP IgG assays have demonstrated good performance compared with established reference methods, while the BLACKBOX® ZIKV ED3 IgG assay has shown high specificity and accuracy, including in the presence of antibodies against other flaviviruses (1–3).

To assess potential cross-reactivity of the the Qualitative Human Marburg Virus Antibody (GP) IgG ELISA kit (MBS109415) from MyBioSource (San Diego, USA), in-house testing was performed in Hamburg, Germany, on 176 plasma or serum samples from blood donors originating from non-endemic regions for MARV (Europe, South America, and Asia). All samples tested negative, except for one sample originating from Colombia. In addition, cross-reactivity with *Orthoebolavirus zaïrense* antibodies was evaluated using 16 plasma samples from well-characterized EVD survivors (4) and no cross-reactivity was observed.

At the time of testing, no MARV-positive serum samples were available for direct assessment of assay performance. However, the limited cross-reactivity observed supported the deployment of the assay for field testing at LFHV-GKD.

## Tables

**Table S1.** Demographic characteristics of study group.

| Characteristic/Site | LFHV-GKD<br>n/N (percent) | LFHV-HRNZE<br>n/N (percent) |
| --- | --- | --- |
| **Median age (IQR), min-max (years)** | 35 (25-50), 2-85 | 36 (19-53), 1-95 |
| **Age group (years)** |  |  |
| <20 | 64/512 (12.5%) | 122/477 (25.6%) |
| 20–35 | 217/512 (42.4%) | 115/477 (24.1%) |
| 36-50 | 111/512 (21.7%) | 110/477 (23.1%) |
| >50 | 119/512 (23.2%) | 129/477 (27.0%) |
| Unknown | 1/512 (0.2%) | 1/477 (0.2%) |
| **Sex** |  |  |
| Female | 291/512 (56.8%) | 234/477 (49.1%) |
| Male | 221/512 (43.2%) | 243/477 (50.9%) |
| **Occupation** |  |  |
| Domestic worker | 185/512 (36.1%) | 120/477 (25.2%) |
| Farming | 79/512 (15.4%) | 57/477 (11.9%) |
| Health worker | 23/512 (4.5%) | 28/477 (5.9%) |
| Other | 132/512 (25.8%) | 132/477 (27.7%) |
| Student* | 70/512 (13.7%) | 134/477 (28.1%) |
| Unknown | 23/512 (4.5%) | 6/477 (1.2%) |
| **Country of origin** |  |  |
| Guinea | 509/512 (99.4%) | 473/477 (99.2%) |
| Other | 3/512 (0.6%) | 4/477 (0.8%) |
| **Region of origin** |  |  |
| N'Zérékoré | 507/512 (99.0%) | 473/477 (99.2%) |
| Other | 5/512 (1.0%) | 4/477 (0.8%) |
| **Prefecture of origin** |  |  |
| Beyla | 0/512 (0.0%) | 34/477 (7.1%) |
| Guéckédou | 488/512 (95.3%) | 1/477 (0.2%) |
| Lola | 0/512 (0.0%) | 23/477 (4.8%) |
| Macenta | 1/512 (0.2%) | 20/477 (4.2%) |
| N'Zérékoré | 2/512 (0.4%) | 387/477 (81.1%) |
| Other | 6/512 (1.2%) | 4/477 (0.9%) |
| Yomou | 15/512 (2.9%) | 8/477 (1.7%) |

IQR: interquartile;
\*Including children in pre-school age (e.g. 3 years old)

**Table S2.** Prevalence of Lassa virus IgG according to demographic factors and samplings years.

| Variable/Site | LASV (IgG) |  |
| --- | --- | --- |
|  | LFHV-GKD<br>Positive/Total (percent) | LFHV-HRNZE<br>Positive/Total (percent) |
| **Sex** |  |  |
| Female | 162/291 (55.7%) | 67/234 (28.6%) |
| Male | 125/221 (56.6%) | 75/243 (30.9%) |
| **Age group (years)** |  |  |
| <20 | 21/64 (32.8%) | 20/122 (16.4%) |
| 20–35 | 103/217 (47.5%) | 41/115 (35.6%) |
| 36-50 | 70/111 (63.1%) | 35/110 (31.8%) |
| >50 | 92/119 (77.3%) | 45/129 (34.9%) |
| Unknown | 1/1 (100.0%) | 1/1 (100.0%) |
| **Occupation** |  |  |
| Domestic worker | 114/185 (61.6%) | 37/120 (30.8%) |
| Farming | 58/79 (73.4%) | 20/57 (35.1%) |
| Health worker | 10/23 (43.5%) | 10/28 (35.7%) |
| Other | 66/132 (50.0%) | 50/132 (37.9%) |
| Student* | 29/70 (41.4%) | 25/134 (18.7%) |
| Unknown | 10/23 (43.5%) | 0/6 (0.0%) |
| **Prefecture of origin** |  |  |
| Beyla | - | 5/4 (14.7%) |
| Guéckédou | 275/488 (56.3%) | 1/1 (100%) |
| Lola | - | 5/23 (21.7%) |
| Macenta | 0/1 (0.0%) | 9/20 (45.0%) |
| N'Zérékoré | 0/2 (0.0%) | 120/387 (31.0%) |
| Other | 0/2 (0.0%) | 0/4 (0.0%) |
| Unknown | 2/4 (50.0%) | - |
| Yomou | 10/15 (66.7%) | 2/8 (25.0%) |
| **Total** | **287/512 (56.0%)** | **142/477 (29.8%)** |

-, no observations; LASV, Lassa virus; IgG, immunoglobulins G.
\*Including children in pre-school age (e.g. 3 years old)

**Table S3.** Prevalence Marburg virus, Zika virus and Crimean-Congo hemorrhagic fever virus (IgG) by demographic characteristics.

| **Variable/ Pathogen** | **MARV (IgG)** | **ZIKV (IgG)** | **CCHFV (IgG)** |
| --- | --- | --- | --- |
|  | **Positive/Total**<br>**(percent)** | **Positive/Total**<br>**(percent)** | **Positive/Total**<br>**(percent)** |
| **Sex** |  |  |  |
| Female | 13/291 (4.5%) | 12/291 (4.1%) | 2/291 (0.7%) |
| Male | 17/221 (7.7%) | 16/221 (7.2%) | 4/221 (1.8%) |
| **Age group (years)** |  |  |  |
| <20 | 4/64 (6.2%) | 4/64 (6.2%) | 0/64 (0.0%) |
| 20–35 | 11/217 (5.1%) | 9/217 (4.1%) | 2/217 (0.9%) |
| 36-50 | 8/111 (7.2%) | 7/111 (6.3%) | 3/111 (2.7%) |
| >50 | 7/119 (5.9%) | 8/119 (6.7%) | 1/119 (0.8%) |
| **Occupation** |  |  |  |
| Domestic worker | 6/185 (3.2%) | 8/185 (4.3%) | 2/185 (1.1%) |
| Farming | 4/79 (5.1 %) | 9/79 (11.4%) | 0/79 (0.0%) |
| Health worker | 2/23 (8.7%) | 1/23 (4.3%) | 0/23 (0.0%) |
| Student* | 10/70 (14.3%) | 3/70 (4.3%) | 1/70 (1.4%) |
| Other / Unknown | 8/155 (5.2%) | 7/155 (4.5%) | 3/155 (1.9%) |
| **Prefecture of origin** |  |  |  |
| Guéckédou | 26/488 (5.3%) | 26/488 (5.3%) | 6/488 (1.2%) |
| Other / Unknown | 4/24 (16.7%) | 2/24 (8.3%) | 0/24 (0.0%) |
| **Total** | **30/512 (5.8%)** | **28/512 (5.4%)** | **6/512 (1.2%)** |

MARV, Marburg virus; ZIKV, Zika virus, CCHFV, Crimea-Congo Hemoragic fever virus; IgG, immunoglobulins G.
\*Including children in pre-school age (e.g. 3 years old)

**Table S4.** Co-seropositivity analysis for one or more pathogens in LFHV-GKD cohort.

| **Number Positive (IgG)**<br>**tests per sample** | **Positive/Total (percent)** |
| --- | --- |
| 0 | 197/512 (38.5%) |
| 1 | 279/512 (54.5%) |
| 2 | 36/512 (7.0%) |
| 3 | 0/512 (0.0%) |
| 4 | 0/512 (0.0%) |

